# The effects of reactive balance training on cardiorespiratory fitness and muscle strength: a pilot randomized controlled trial

**DOI:** 10.1101/2024.12.16.24319109

**Authors:** Augustine J. Devasahayam, Azadeh Barzideh, David Jagroop, Cynthia Danells, Elizabeth L. Inness, Susan Marzolini, Sunitha Mathur, Paul Oh, Avril Mansfield

**Author notes:** **Corresponding author:** Avril Mansfield, 550 University Ave, Toronto, ON, M5G 2A2, Canada.

## Abstract

**Background:** Reactive balance training (RBT) may improve multiple components of physical fitness, including aerobic capacity and muscle strength. However, there have been no studies examining its effects on these factors in people with stroke.

**Objectives:** The objectives of this pilot study were to determine the feasibility of a non-inferiority randomized controlled trial, comparing aerobic and strength training (AST) and RBT, considering sample size (primary objective), rates of accrual and withdrawal, intervention adherence, missing data, preliminary effects, and harms (secondary objectives).

**Methods:** People who were at least six months’ post-stroke and could stand independently for >30 seconds were recruited. Peak oxygen consumption 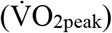 was measured by cardiopulmonary exercise test. Peak isokinetic torques for knee extension and flexion were measured by dynamometer.

**Results:** Twenty-three participants (6 women) were randomized into AST and RBT groups. Four-hundred participants per group were estimated to be required for the main trial considering 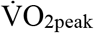 and peak isokinetic torque as primary outcomes. Rates of accrual and withdrawal were 2 participants for every quarter and 30%, respectively. On average, AST participants attended 29.6/36 sessions (range: 18-36) and RBT participants attended 23.5/36 sessions (range: 1-35). Data were missing for 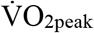 (n=2) and ABC scale (n=1) as participants declined testing. 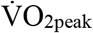 and peak knee extension torque of more-affected legs improved post-intervention in both groups. Ten adverse events related to study interventions resolved without medical attention.

**Conclusion:** Progressing to a definitive single-site trial is not feasible given the large required sample size, low accrual, and high withdrawal rates.

**Trial registration:** NCT04042961

## INTRODUCTION

Stroke is a leading cause of long-term adult disability in Canada.^1^ The number of Canadians living with the effects of stroke is projected to increase from 405,000 in 2013 to up to 726,000 by 2038, reflecting a significant public health burden.^1^ After stroke, patients admitted to acute inpatient care are either discharged to their homes independently, discharged home with home care services, transferred to an inpatient rehabilitation program, transferred to long-term or complex continuing care, or to another acute care facility.^2^ These transitions in stroke care are directed in part by the patients’ physical,^3^ cognitive,^4^ and social ^5^ capacities that are related to fall risk. Between 1 and 6 months after a stroke, up to 37% of people with stroke experience falls, and this increases to as much as 73% within a year post-stroke.^6,7^

Balance is an essential component of physical fitness that determines transitions in stroke care.^8-10^ A person with stroke who does not have the ability to recover from a loss of balance is at risk of sustaining fall-related injuries,^11^ which in turn may lead to further disability, death, and increase in health care costs.^1^ Reactive balance training (RBT) is a type of exercise that improves reactions to the sudden loss of balance,^12^ and may decrease the risk of falls after discharge from inpatient rehabilitation.^13,14^ As RBT involves evoking whole-body balance reactions repeatedly, it is possible that multiple components of physical fitness could improve, including aerobic capacity and lower extremity muscle strength.^15^ To our knowledge, no studies have investigated the effect of RBT in improving these components of physical fitness among people living with stroke.

We aimed to determine the effect of RBT on cardiorespiratory fitness and lower extremity isokinetic muscle strength in people with chronic stroke compared to aerobic and strength training (AST), through a definitive non-inferiority randomized controlled trial.^16^ The secondary aims were to determine the effect of RBT and AST on balance control and balance confidence.^16^

In this paper, we report the interim results from a pre-planned internal pilot study where we planned to re-estimate the target sample size for the non-inferiority randomized controlled (main) trial using the variance of the primary outcomes measured in the internal pilot study.^16^ The primary objective of this pilot study was to determine the revised target sample size for the main trial. The secondary objectives were to assess: (1) the feasibility of recruiting the revised target sample size for the main trial (rates of accrual and withdrawal from the pilot study); and (2) the feasibility of proceeding with the main trial considering the revised target sample size (intervention adherence, proportion of missing data, preliminary training effects, and harms related to the study interventions in the pilot study).

## 2. Methods

### Trial design

This was an assessor-blinded pilot randomized controlled trial. The study was approved by the research ethics boards of the University Health Network, Toronto, Ontario (protocol number: 18-5784) and the University of Toronto (protocol number: 37859). The study was registered with clinicaltrials.gov prior to recruiting the first participant (registration number: NCT04042961) and the full study protocol is available elsewhere.^16^ The study is reported according to the CONSORT guidelines for reporting non-pharmacologic treatments,^17^ and pilot studies,^18^ and the CONSERVE statement for reporting trials modified due to the coronavirus disease 2019 (COVID-19) pandemic.^19^

This study was impacted by the onset of the COVID-19 pandemic in the following ways: complete research shut-down and research staff re-deployment, which interrupted study activities for 12 months total from March 2020 to December 2020, and from January 2022 to February 2022; partial loss of funding due to financial constraints from our funder; and participant withdrawal and recruitment challenges due to potential participants not wanting to risk COVID-19 exposure.

### Participants

Participants were recruited from December 2019 to January 2024 by: (1) inviting individuals from previous studies conducted by the investigators; (2) seeking referrals from stroke rehabilitation programs at the Toronto Rehabilitation Institute; (3) using research participant databases at the Toronto Rehabilitation Institute; and (4) placing advertisements in the community, including health centres and local newspapers.^16^ Inclusion criteria were: (1) diagnosis of chronic stroke (>6 months post-stroke); (2) age ≥ 20 years; (3) living in the community; (4) able to stand independently without upper-limb support for >30 seconds; and (5) able to tolerate at least 10 postural perturbations while wearing a safety harness. The following exclusion criteria were applied: (1) height taller than 2.1m and/or body mass more than 150kg (due to limits of the safety harness system); (2) presence of other neurological conditions affecting balance control (e.g., Parkinson’s disease); (3) history of lower extremity amputation; (4) cognitive, language, or communication impairments affecting understanding of instructions; (5) recent significant illness, injury, or surgery within the last 6 months; (6) diagnosis of severe osteoporosis with fracture; (7) uncontrolled hypertension; (8) uncontrolled diabetes or on insulin therapy; (9) contraindications to exercise testing, such as symptomatic aortic stenosis, complex life-threatening arrhythmias, unstable angina, or orthostatic blood pressure decrease of >20 mmHg with symptoms; (10) acute or chronic illness or injury likely to be exacerbated by exercise (e.g., recent lower-extremity fracture); (11) currently undergoing in-or out-patient physiotherapy involving aerobic training, balance training, or lower limb strength training; (12) participation in regular exercise that met the recommended physical activity guidelines (at least 150 minutes of moderate-to-vigorous or at least 75 minutes of vigorous physical activity per week) as assessed using the moderate and vigorous components of the Leisure Time Exercise Questionnaire (LTEQ) in the month prior to study initiation; and/or (13) receipt of RBT within the past year. Written informed consent was obtained from all participants.

### Interventions

Interventions were administered by a physiotherapist. Participants completed three 60-minute exercise sessions per week for 12 weeks. Participants in the AST group performed 30 minutes of aerobic training and 30 minutes of strength training. The aerobic training prescription was derived from a combination of various methods: anaerobic threshold (V-slope and the Ventilatory Equivalence method); 60-80% of heart rate reserve, 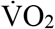 reserve, and peak oxygen consumption 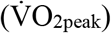. Aerobic training involved treadmill walking, recumbent stepping, or upright cycling chosen based on the participants’ mobility impairment and modality preference. Each session included a five-minute warm-up, progressive exercise up to 20 minutes (based on prescribed target heart rates and/or a rating of perceived exertion (RPE) of 11-16 on the Borg 6-20 scale^20^), and five-minute cool-down. Strength training involved 2 sets of 8 exercises per session: (8-10 repetitions) lunge, squat, heel raise, ankle dorsiflexion, hip flexion, knee extension and flexion, abdominal curl-up, wall push up, bicep curl, and triceps extension. Resistance was 50-60% of 1 repetition maximum for the first 2 weeks and 70% of 1 repetition maximum thereafter; 1 repetition maximum was assessed at weeks 1, 3, 6, and 10.

Participants in the RBT group performed individualized RBT that was designed to target participants’ specific impairments. Each session included a five-minute warm-up, exercises consisting of at least 60 external and internal perturbations,^21^ and a five-minute cool-down. During each session, the difficulty of the prescribed tasks was targeted in such a way that participants would “fail” to recover balance (i.e., use an upper extremity recovery response, a multistep (>2 steps) recovery response, or require assistance from the PT or harness) at least 50% of the time.

### Randomization and blinded allocation

Participants were assigned using blocked stratified randomization with allocation concealment to one of two training groups: AST or RBT. There were 4 strata based on 2 factors, i.e., Berg Balance Scale score (two levels: 46 vs >46), and baseline 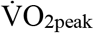 (two levels: 15 ml/kg/min vs ≥15 ml/kg/min).

The random allocation sequence was computer-generated. At baseline, the outcome measures were administered by a research assistant who was blinded to group allocation. After the baseline assessment, information required for randomization (Berg Balance Scale score and 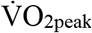), and information pertaining to participant eligibility was communicated by the research assistant to the principal investigator. Group allocation was performed independently by the principal investigator who was not involved in recruiting, outcome measure assessment, or administering the interventions. After randomization, the principal investigator communicated the group allocation to the physiotherapist who administered the interventions.

### Outcomes

#### Revised target sample size

According to the plan established *a priori*, we aimed to recruit and allocate at least 20 participants for the internal pilot study.^16^ The revised target sample size for the main trial was then calculated using the variance of five outcome measures from this pilot study (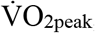, and four lower extremity isometric strength measures, i.e., maximal torque value for knee extension and flexion of the more and less affected lower extremities).^16^ We calculated individual sample sizes for each outcome and selected the largest sample size requirement as the revised target sample size for the main trial to ensure adequate power for each test.^22^

The target sample size for the main trial was calculated using the following formula:^23,24^

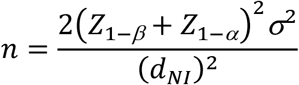

where n is the number of participants per group, Z_1-β_ is the Z statistic for the probability of a type II error (β=0.1), Z_1-α_ is the Z statistic for the probability of a type I error (α=0.05), σ^2^ is the estimated population variance for the outcomes, and d_NI_ is the non-inferiority margin. We defined the non-inferiority margin using the standard error of measurement (SEM) from previous studies on test-retest reliability of aerobic capacity^25^ and lower-extremity strength^26^ post-stroke.

#### Feasibility outcomes

The feasibility of recruiting the revised target sample size for the main trial was determined by: (1) the rate of accrual into the pilot study during recruitment period, and (2) the rate of withdrawal from the pilot study after randomization into study groups. Rate of accrual was assessed from the number of participants recruited every quarter (3 months) during the active recruitment periods from December 2019 to January 2024 (i.e., excluding periods when recruiting was paused due to the COVID-19 pandemic). Rate of withdrawal was assessed from the number of participants who withdrew from the pilot study after randomization into study groups.

The feasibility of proceeding with the main trial considering the revised target sample size was determined by: (1) intervention adherence, (2) proportion of missing data, (3) preliminary training effects, and (4) harms related to the study interventions observed during the pilot study. Intervention adherence was assessed by determining the extent to which the participants completed the prescribed exercises, i.e., by counting the duration of aerobic exercise, number of strength exercises, number of perturbations, and the total number of sessions completed by the participants. Proportion of missing data was calculated for each outcome measure.

The preliminary training effects were assessed through the following outcome measures: 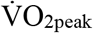, lower extremity muscle strength, anticipatory balance control, reactive balance control, balance confidence, walking endurance, and functional lower extremity strength.

A symptom-limited cardiopulmonary exercise test (CPET) was conducted to determine 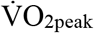 ml/kg/min using either a semi-recumbent cycle ergometer with specialized pedals to support the feet (Ergoline, Ergoselect 1000, Blitz, Germany), an upright cycle (Ergoselect 200P, Germany), or a treadmill, depending on the individual’s balance and ability to control their leg/foot position in pedals. A treadmill was used if participants met at least one of the following criteria during the initial assessment session: 1) Chedoke-McMaster Stroke Assessment (CMSA)^27^ leg and foot scores ≥4; 2) 6-minute walk test (6MWT) ≥144m;^28^ and 3) capability to walk on the treadmill for at least three minutes at 2.7 km·h^−1^ with a 10% incline (i.e. the first stage of the Standard Bruce exercise test protocol).^29^ Participants not meeting these criteria used either semi-recumbent stepper or upright cycle based on the participants’ mobility impairment and modality preference. Breath-by-breath gas samples were collected via calibrated metabolic cart (SensorMedics Vmax Encore, San Diego, California, USA). Twelve-lead electrocardiogram, rating of perceived exertion (RPE),^20^ and blood pressure were monitored throughout the test.

Peak isokinetic torque was measured to determine maximal lower extremity muscle strength using a Biodex System 4 Pro dynamometer (Biodex Medical Systems, New York, USA). Participants were seated with their hips and knees at 90· of flexion, and the dynamometer was aligned with lateral femoral condyle of the tested knee joint. To secure the participant in place for testing, straps were placed across the shoulders, waist, and the thigh of the tested leg. Each participant was asked to perform maximal voluntary contractions (MVCs) on their less affected leg first, at an angular velocity of 60· per second for both knee extension and flexion. To ensure accurate and consistent measurements, participants were given standardized instructions and encouraged to exert maximal effort during each trial. They performed a series of warm-up, submaximal concentric contractions (3 repetitions) followed by five isometric MVCs for both knee extension and flexion, with a 60-second rest period between each trial to minimize fatigue. Isokinetic torque was measured between 20· and 90· of knee flexion. For the eccentric testing conditions, torque limits of 150, 200, and 250 Nm were selected based on the initial concentric MVC test (i.e. if participants exceeded 200 Nm in the first condition, then a torque limit of 250 Nm was selected for the proceeding eccentric tests of that tested leg). All measurements were taken for both the more and less affected lower extremities. A single maximal torque value for knee extension and flexion of the more and less affected lower extremities was obtained by calculating the average of the three highest achieved across five maximal trials.^16^

Functional balance was assessed using the Berg balance scale (BBS)^30^ and mini-Balance Evaluation Systems Test (mini-BEST),^31^ and balance confidence was assessed using the Activities-specific Balance Confidence (ABC) scale.^32^ The BBS is a 14-item observational rating scale that provides a measure of functional static and dynamic balance. The mini-BEST is a 14-item observational rating scale that assesses systems underlying balance control, including reactive balance control and dynamic stability during walking.^31^ The ABC scale^32^ is a questionnaire used to assess balance confidence during daily activities before and after training, asking participants to rate, on a scale from 0-100%, how confident they would be performing 16 everyday tasks, without feeling unsteady, losing their balance or falling.

The six-minute walk test (6MWT) was conducted to assess functional walking endurance.^33^ The 6MWT is a self-paced, exercise test that measures the maximum distance a person can walk in 6 minutes. Participants were asked to walk back and forth along the 30-metre path for 6 minutes, covering as much distance as possible; the total distance walked was recorded. The 30-second sit-to-stand (STS (30s)) test was conducted to assess the functional walking strength.^34^ Participants were asked to sit on a chair with a height of 43.2 cm, placed against a wall for stability. They were asked to rise to a full stand and return to a fully seated position as many times as possible for 30 seconds and the total number of complete sit-to-stand repetitions was recorded.

Harms related to the training interventions were assessed by recording the number and type of adverse events reported by the participants during and after completing training sessions in the respective session log and by determining whether they were possibly, probably, or definitely related to study interventions by the physiotherapist who conducted the training sessions and the principal investigator.

### Data analysis

All data are presented as mean (standard deviation) or with 95% confidence intervals, unless stated otherwise.

Rate of participant accrual throughout the recruitment phase of this pilot study was determined by averaging the number of participants recruited every three months, while excluding periods when recruitment was paused due to the COVID-19 pandemic. Rate of withdrawal was calculated by dividing the total number of participants who dropped out of the study by the total number of participants who were randomized into the study groups.

Intervention adherence was calculated by taking an average of the total number of sessions attended by the participants out of the 36 possible training sessions for AST and RBT groups separately. An average of the individual components of the training sessions attended was calculated by counting the minutes of aerobic exercise and number (repetitions and sets) of strength exercises completed per session. A count of the number of perturbations received were recorded to calculate the average number of perturbations per session and for all RBT sessions combined.

The proportion of missing data was calculated for each outcome measure separately. To calculate the proportion of missing data, the total number of participants who declined or were unable to complete the key outcome measures was divided by the total number of participants who successfully completed the data collection.

The magnitude of training difference in 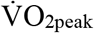, peak muscle strength of knee extensors and flexors for both more and less affected sides, BBS score, mini-BEST score, ABC scale score (%), 6MWT distance (m), 30-s sit-to-stand (number) score was calculated by computing the post-to pre-intervention difference for each participant, and the mean and 95% confidence intervals of the differences for each group were calculated. The number of adverse events related to study interventions was counted.

## RESULTS

### Recruitment

Recruitment occurred between 3 December 2019 and 23 January 2024. Recruitment of new participants was paused between March 2020 and December 2020, and between January 2022 and February 2022 due to the COVID-19 pandemic. Twenty-eight participants were enrolled in the study, of which 12 were allocated to the AST group and 11 were allocated to the RBT group (Figure 1). Participant characteristics are in Table 1.

**Table 1:**
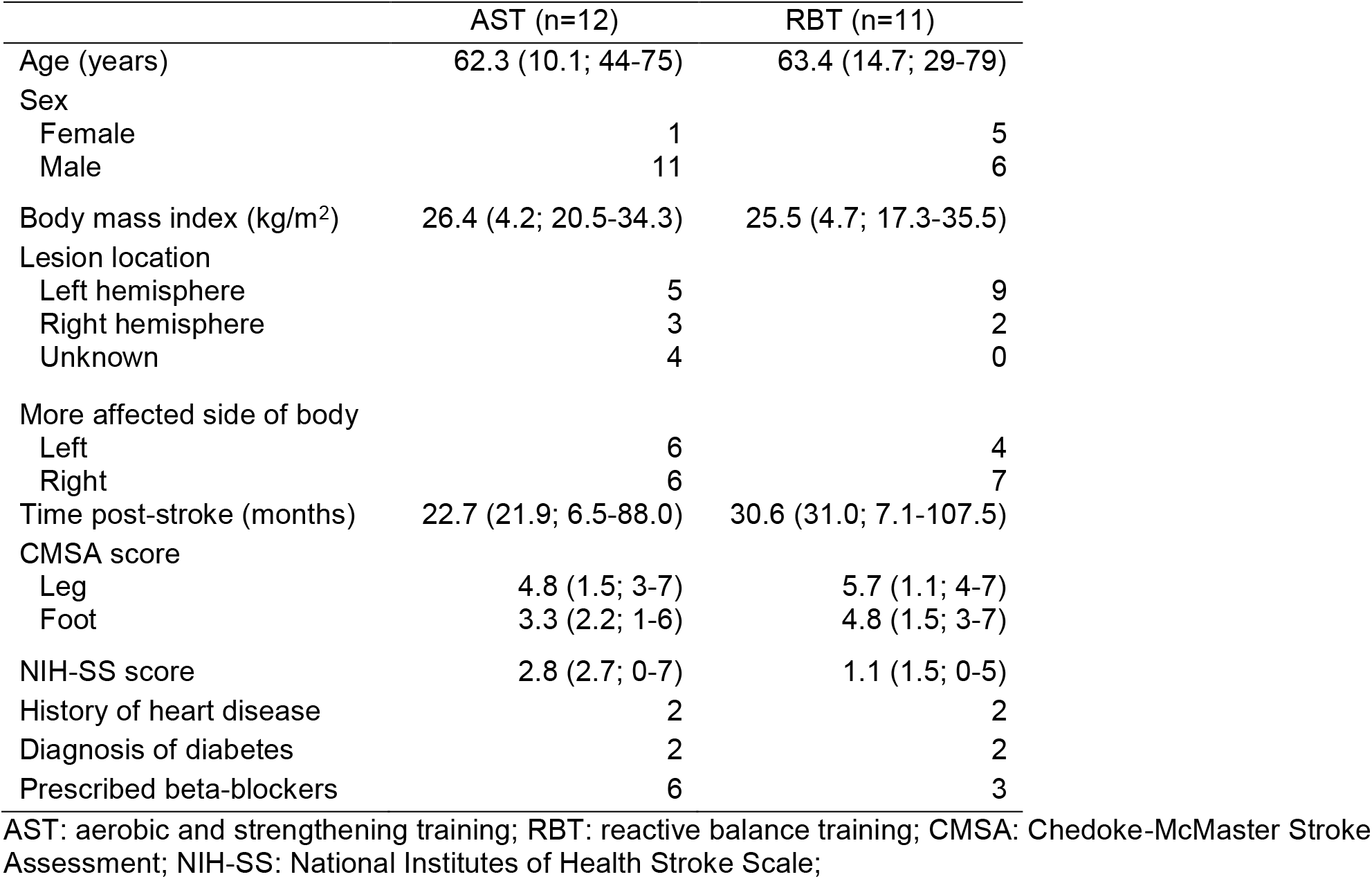
Participant characteristics. Values presented are means with standard deviations and ranges in parentheses for continuous variables, or counts for categorical variables.

|  | AST (n=12) | RBT (n=11) |
| --- | --- | --- |
| Age (years) | 62.3 (10.1; 44-75) | 63.4 (14.7; 29-79) |
| Sex |  |  |
| Female | 1 | 5 |
| Male | 11 | 6 |
| Body mass index (kg/m<sup>2</sup>) | 26.4 (4.2; 20.5-34.3) | 25.5 (4.7; 17.3-35.5) |
| Lesion location |  |  |
| Left hemisphere | 5 | 9 |
| Right hemisphere | 3 | 2 |
| Unknown | 4 | 0 |
| More affected side of body |  |  |
| Left | 6 | 4 |
| Right | 6 | 7 |
| Time post-stroke (months) | 22.7 (21.9; 6.5-88.0) | 30.6 (31.0; 7.1-107.5) |
| CMSA score |  |  |
| Leg | 4.8 (1.5; 3-7) | 5.7 (1.1; 4-7) |
| Foot | 3.3 (2.2; 1-6) | 4.8 (1.5; 3-7) |
| NIH-SS score | 2.8 (2.7; 0-7) | 1.1 (1.5; 0-5) |
| History of heart disease | 2 | 2 |
| Diagnosis of diabetes | 2 | 2 |
| Prescribed beta-blockers | 6 | 3 |

AST: aerobic and strengthening training; RBT: reactive balance training; CMSA: Chedoke-McMaster Stroke Assessment; NIH-SS: National Institutes of Health Stroke Scale;

**Figure 1:**
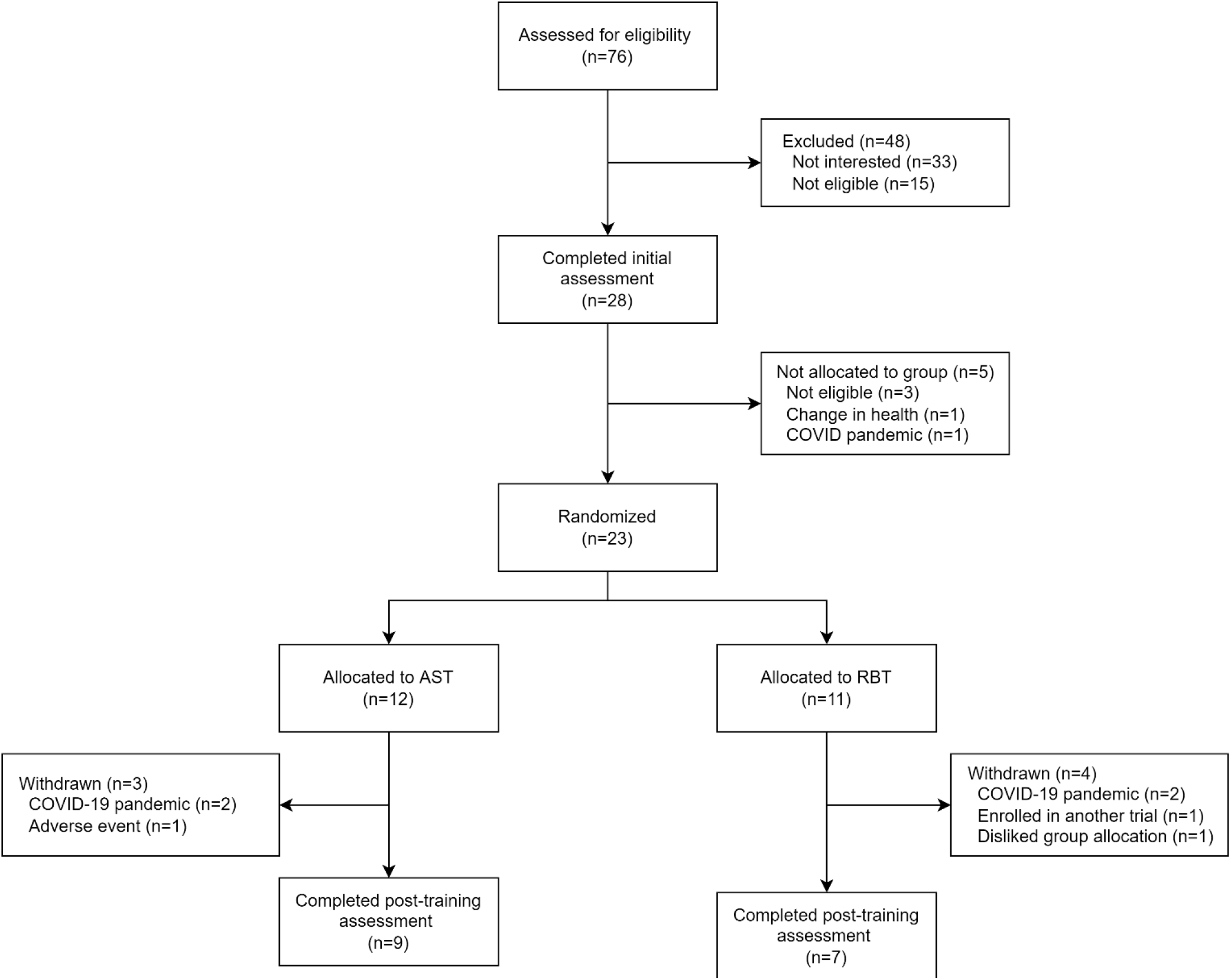
Participant recruitment flow.

### Revised target sample size for the main trial

To achieve the desired statistical power to show non-inferiority of the RBT group compared to the AST group for 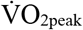 and peak isometric strength, while accommodating a 30% anticipated withdrawal rate, it is estimated that 400 participants per group are required for the main trial. The variances, SEMs, and associated sample sizes for each of the primary outcomes are presented in Table 2.

**Table 2:** Sample size estimation for the main trial. The non-inferiority margin is the SEM. The sample size estimate is the number of participants required per group after accounting for 30% withdrawal rate.

| Outcome measure | Variance | SEM | Sample size estimate |
| --- | --- | --- | --- |
| $\dot{V}O_{2\text{peak}}$ (ml/kg/min) | 15 | 1.0 | 400 |
| More affected limb knee extension strength (Nm) | 330 | 12.4 | 57 |
| More affected limb knee flexion strength (Nm) | 114 | 10.1 | 30 |
| Less affected limb knee extension strength (Nm) | 510 | 10.6 | 121 |
| Less affected limb knee flexion strength (Nm) | 293 | 7.4 | 143 |

$\dot{V}O_{2\text{peak}}$ : peak oxygen consumption; ml: milli-liter; kg: kilogram; min: minute; Nm: newton-meter; SEM: standard error of measurement;

### Rates of accrual and withdrawal

Twenty-eight participants were recruited over 38 months during the active recruitment periods from December 2019 to January 2024. The mean rate of accrual was 2 participants per quarter.

Seven participants withdrew from the study after allocation (3/12 AST, 4/11 RBT); therefore, outcome data were available for 9 AST and 7 RBT participants (Figure 1).

### Intervention adherence

AST participants attended a mean of 29.6 sessions (range: 18-36 sessions), with 8/12 (67%) participants attending at least 30/36 sessions. AST participants completed all prescribed strength exercises for an average of 48.4% of sessions attended (standard deviation: 32.7% of sessions attended; range: 0.7-95% of sessions attended), and completed at least 20 minutes of aerobic exercise for an average of 52.6% of sessions (standard deviation: 25.4% of sessions attended; range: 0-87.1% of sessions attended). For all AST sessions combined, the mean rate of perceived exertion was 11.6 (out of 20; standard deviation: 1.3). One RBT participant withdrew from the study following randomization but prior to attending any training sessions. The remaining 10 RBT participants attended a mean of 23.5 sessions (range: 1-35 sessions), with 3/10 participants attending at least 30/36 sessions. Participants experienced an average of 1418 perturbations during all sessions combined, which corresponded to an average of 58.3 perturbations/session (standard deviation: 27.8 perturbations/session; range 0-105 perturbations/session). For all RBT sessions combined, the mean rate of balance recovery failures was 28.9% (standard deviation: 16.7%; range: 0-59.4%) and mean rate of perceived challenge was 2.4 (out of 5; standard deviation: 0.6).

### Missing data

Data were missing for two outcomes due to participants declining testing (Table 3). The rate of missing data was 1/9 and 1/7 for 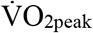 in AST and RBT groups respectively, and 1/9 for the ABC scale in the AST group (Table 3).

**Table 3:** Training difference. Values presented are means with standard deviations in parentheses at each assessment time, and means with 95% confidence intervals in brackets for the pre-to-post training difference. The pre to post training difference was calculated as post-training minus pre-training; therefore, a positive value indicates an improvement with training.

|  | AST (n=9) |  |  | RBT (n=7) |  |  |
| --- | --- | --- | --- | --- | --- | --- |
|  | Pre-training | Post-training | Difference | Pre-training | Post-training | Difference |
| **Outcome measures** |  |  |  |  |  |  |
| $\dot{V}O_{2peak}$ (ml/kg/min)* | 15.69 (2.92) | 17.35 (3.58) | 1.67 [0.21, 3.12] | 16.70 (6.25) | 19.53 (8.31) | 2.83 [1.37, 4.29] |
| More affected knee extension strength (Nm) | 46.9 (21.1) | 49.0 (20.5) | 2.2 [-0.3, 4.6] | 54.5 (9.4) | 63.7 (24.9) | 9.2 [5.7, 12.7] |
| More affected knee flexion strength (Nm) | 24.9 (19.0) | 24.5 (18.7) | -0.4 [-2.8, 2.0] | 26.5 (7.3) | 26.1 (4.5) | -0.3 [-2.2, 1.5] |
| Less affected knee extension strength (Nm) | 106.1 (44.6) | 114.8 (42.1) | 8.7 [5.4, 12.0] | 88.0 (17.4) | 81.7 (25.0) | -6.3 [-9.2, -3.4] |
| Less affected knee flexion strength (Nm) | 51.6 (25.9) | 54.2 (18.4) | 2.6 [0.1, 5.1] | 44.4 (16.8) | 31.2 (10.7) | -13.1 [-16.2, -10.1] |
| Berg balance scale (score) | 46.7 (6.0) | 49.0 (4.8) | 2.3 [0.7, 4.0] | 49.9 (5.8) | 52.0 (4.4) | 2.1 [1.0, 3.3] |
| Mini-BEST (score) | 20.8 (2.7) | 19.8 (3.4) | -1.0 [-2.0, 0] | 22.9 (1.3) | 22.9 (4.2) | 0 [-1.3, 1.3] |
| ABC (%)† | 65.8 (9.0) | 71.1 (15.2) | 5.3 [2.9, 7.7] | 76.7 (14.7) | 80.9 (13.7) | 4.3 [2.8, 5.7] |
| 6MWT (m) | 271.6 (138.9) | 300.3 (145.0) | 28.8 [25.0, 32.6] | 374.6 (102.4) | 392.6 (114.5) | 18.0 [14.2, 21.8] |
| 30-s sit-to-stand (number) | 8.0 (3.0) | 10.0 (2.6) | 2.0 [1.1, 2.9] | 12.9 (4.2) | 15.9 (6.4) | 3.0 [1.6, 4.4] |

\*n=8 for the AST group and n=6 for the RBT group, and †n=8 for the AST group participants declining the test; AST: aerobic and strengthening training; RBT: reactive balance training; $\dot{V}O_{2peak}$ : peak oxygen consumption; ml: milli-liter; kg: kilogram; min: minute; Nm: newton-meter; BEST=Balance Evaluation Systems Test; ABC: Activities-specific Balance Confidence; 6MWT: 6-minute walk test, m: meter; s: second;

### Training difference

The pre to post difference after AST and RBT in individual participants are presented in Figure 2 and Table 3. On average, 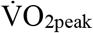 improved for both groups pre to post intervention (Figure 2a and 2b). The mean peak knee extension torque pre to post intervention for the more-affected leg increased in both groups (Figure 2c and 2d). However, the mean peak knee flexion torque pre to post intervention for the more-affected leg decreased in both groups (Figure 2e and 2f). The mean peak knee extension and knee flexion torques pre to post intervention for the less-affected leg increased in the AST group and decreased in the RBT group (Figure 2g, 2h, 2i, and 2j).

**Figure 2:**
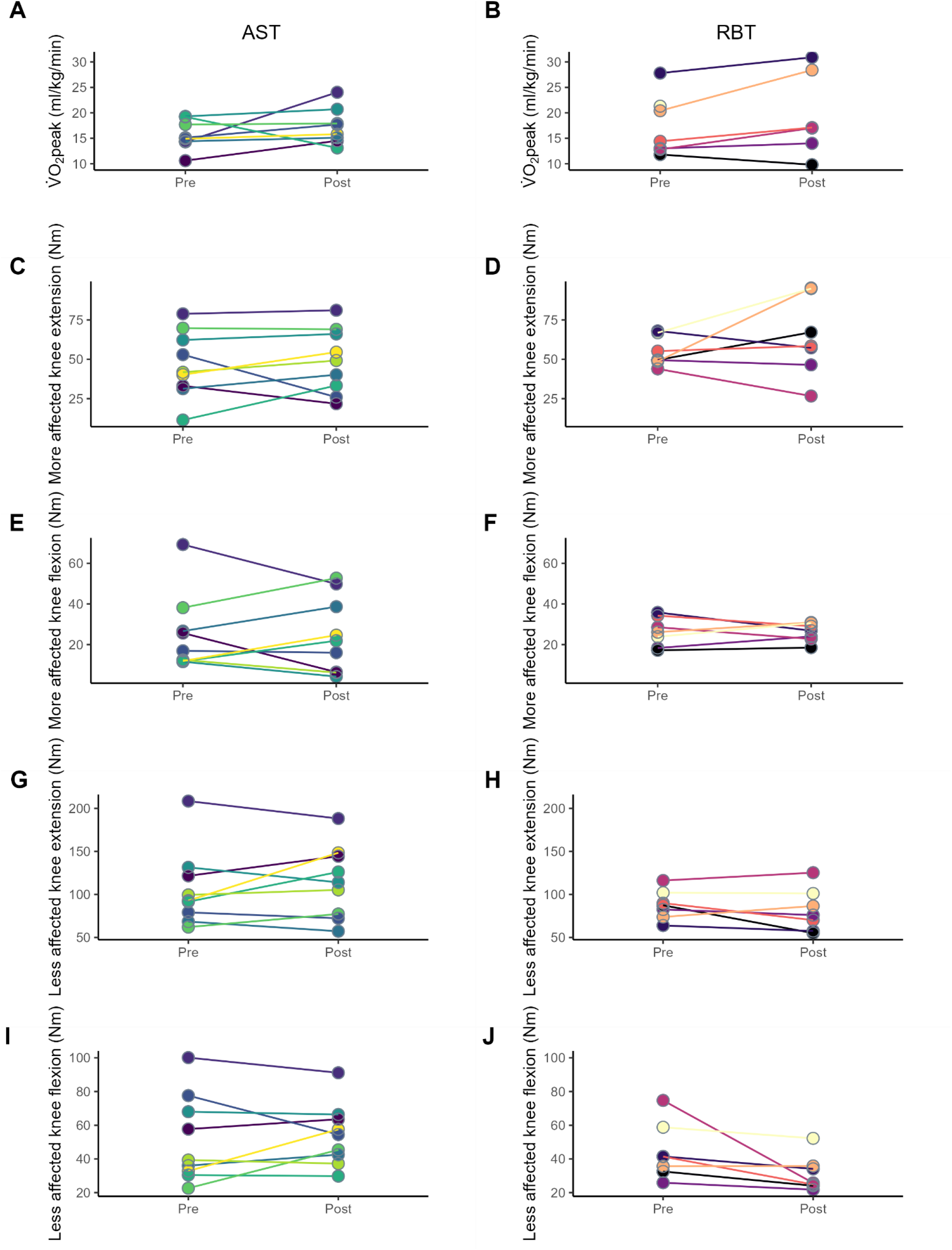
Pre to post difference after aerobic and strength training and reactive balance training. Each data point represents an individual participant.

The mean BBS score, ABC scale scores, 6MWT distance, and STS repetitions increased in both groups post intervention. The mean mini-BEST scale scores decreased in the AST group and did not change in the RBT group post intervention.

### Harms

Ten adverse events were possibly, probably, or definitely related to study interventions among the 23 randomized participants. Events were: lower extremity pain (2 AST, 1 RBT), shoulder pain (1 AST), back pain (1 AST, 1 RBT), minor injury (2 RBT), hyperhidrosis (1 AST), and a fall after training session that was possibly related to fatigue (1 RBT). All adverse events were considered minor as they resolved without medical intervention, although the participant with hyperhidrosis withdrew from the study due to this adverse event.

## DISCUSSION

This is a report from an internal pilot study of a randomized controlled (main) trial. Our pilot study indicates that, due to the large sample size required, low accrual rates, and high withdrawal rates, progressing to a definitive randomized controlled trial is not feasible. However, the results suggest that

RBT may be an effective and safe intervention for improving 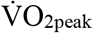 and functional capacity in patients with chronic stroke. Future research should explore alternative trial designs, refine recruitment strategies, and consider multi-site studies to increase sample size and improve generalizability of results.

We initially estimated that 35 participants per group would be needed for the main trial, accounting for a 20% withdrawal rate, based on variance from a previous RCT.^16^ We also planned to re-estimate the target sample size for the main trial using variance from the primary outcomes measured in this internal pilot study, with the main trial designed as a non-inferiority RCT.^16^ As planned a priori, we recruited at least 20 participants for random allocation into two study groups in this pilot study.^16^ The variance was larger for all primary outcomes measured in this pilot study (Table 2) when compared to the variance we used from the previous RCT to initially estimate the sample size needed for the main trial.^16^ We initially based our estimate of variance in 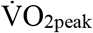 on a large RCT that had recruited similar participants to ours (i.e., with chronic stroke).^35^ However, this study reported very low variance in 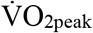 compared to other trials;^36^ the variance in change in 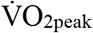 with training among stroke participants generally ranges widely, from 6.5 to 52.2 ml/kg/min.^37-41^ The higher than expected variances in this pilot study suggests that the initial sample size was underestimated, resulting in a need for a larger sample size (n=400) to achieve adequate statistical power for the main trial. In our context, the large sample size requirement is largely due to the non-inferiority design, which requires sufficient statistical power to demonstrate that the experimental RBT intervention is not worse than the control AST intervention by more than a pre-specified non-inferiority margin.^42^ Additionally, low accrual rates and high withdrawal rates further contributed to the increased sample size needed.

In our pilot study, the rate of accrual was 2 participants for every 3 months (0.74 participants per month) during the active recruitment period from December 2019 to January 2024. The overall withdrawal rate was 7/23 for both groups combined. Challenges with recruiting and achieving the target sample size are common for stroke rehabilitation trials, with ∼40% of published trials recruiting <90% of the target sample size.^43^ However, recruiting challenges are likely even more prevalent as failure to recruit an adequate sample size is significant predictor of non-publication of clinical trial results.^44^ Among over 500 stroke rehabilitation RCTs, the rate of accrual is, on average 1-2 participants per month per site.^45^ The low accrual and high withdrawal rates in our pilot study were possibly due to the public health restrictions in place for COVID-19 pandemic. However, even at the end of the study when pandemic restrictions were lifted, our recruitment rate was still <1 participant/month. A previous RBT trial at our site with similar eligibility criteria recruited 3-4 participants/month.^46^ The intervention volume was much lower in the previous trial (2 sessions per week for 6 weeks vs 3 sessions per week for 12 weeks in the current trial). The increased time requirement, and consequently the increased burden on participants, may have impacted our ability to recruit participants into this trial.^47^

In this study, the mean rate of adherence to intervention was 82% for AST participants and 65% for RBT participants. Reasons for non-attendance included COVID-related cancellations, winter weather and long commutes, schedule conflicts such as participant vacations and medical appointments, and illness. Therefore, we believe our adherence rates are comparable to those in previous studies and may demonstrate the feasibility of conducting similar RBT interventions in future studies. Previous studies on RBT in chronic stroke have reported higher adherence rates, ranging from 87-100%.^46,48,49^ However, these comparisons may not be entirely appropriate due to the increased duration of the RBT intervention (12 weeks) compared to previous studies (3-10 weeks). We used a longer duration of RBT intervention in our study to match the duration of the AST intervention; improvements in cardiorespiratory fitness and muscle strength typically take 10-12 weeks. ^50,51^ However, such a long duration of RBT is likely unnecessary for improving reactive balance control,^52^ as many participants reached a plateau long before the end of the program. Consequently, we found that it was often difficult to progress the magnitude of the manual perturbations to sufficiently challenge participants’ reactive balance control toward the end of the program. Therefore, there may be limited value in implementing such a long duration of RBT in future studies.

The mean change in 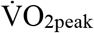 from pre to post intervention was, on average, >1.6 mL/kg/min for the AST group and >2.8 ml/kg/min for the RBT group. The minimal clinically important difference in 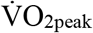 for patients with chronic stroke following a rehabilitation intervention has not yet been determined.^53,54^ Systematic reviews report improvements in 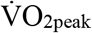 for people with chronic stroke of 1.4-2.27 ml/kg/min for multiple types of exercise^36,55^ or 3.51 ml/kg/min for cardiorespiratory exercise only.^36^ The improvement in 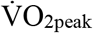 observed in the RBT group in our study is consistent with the findings of these previous studies. Keteyian et al. reported that every 1.0 ml/kg/min increase in 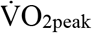 was associated with an approximate 15% decrease in risk of mortality among people with coronary heart disease.^53^ Therefore, the mean improvement in 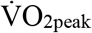 observed in our study may be clinically meaningful. The change in STS (30s) pre to post intervention was, on average, 2.0 or more repetitions in both groups. There is no existing data on either minimal or major clinically important difference of the STS (30s) test for individuals with chronic stroke. However, a cut-off score of at least 2 or more repetitions in the STS (30s) test was considered to provide a reasonably reliable and valid indicator of clinically important improvement at the group level in older adults (41-85 years) with osteoarthritis.^56^ The clinically significant changes seen in these two key outcomes in this pilot study may become non-significant with a larger sample size and sufficient statistical power in the main trial, while other important outcomes may emerge as meaningful. Additionally, at the individual participant level, using clinically important thresholds may incorrectly classify participants with scores below the mean as not having experienced a clinically meaningful change, even if they actually have.

### Limitations

We aimed to recruit and allocate at least 20 participants into two study groups in total, however only 16 completed the 12-week intervention. We acknowledge that the smaller sample size of 16, which resulted from recruitment challenges and non-completion of intervention, may not have been enough to estimate the revised target sample size for the main trial. Furthermore, the feasibility of recruiting the revised target sample size for the main trial was determined by the accrual and withdrawal rates observed during the pilot study. Factors like varying recruitment rates, changes in funding, and increased competition from other trials at the research site after the pandemic could have affected this assessment. These factors may limit the applicability of the pilot study’s findings to the main trial and highlight areas where further amendments to the study protocol may be needed for future trials.

## Conclusions

In this pilot study, the large variance in the primary outcomes led to an increased sample size requirement (n=400 per group) for the main trial. Furthermore, the rate of accrual was slow, with only 2 participants recruited every 3 months. Additionally, participant retention was a challenge, as 7 participants discontinued the study after allocation, 3 from the AST group (25%) and 4 from the RBT group (36%). While AST participants showed good adherence to intervention, attending an average of 29.6/36 sessions, with 67% attending ≥30 sessions, RBT participants had comparably lower adherence, attending 23.5/36 sessions on average, with 30% attending ≥30 sessions. Given the large required sample size, low accrual rates, and high withdrawal rates, progressing to a single-site definitive randomized controlled trial is not feasible.

## Data Availability

All data produced in the present work are contained in the manuscript.

## ACKNOWLEDGEMENTS

This study was funded by the Heart and Stroke Foundation of Canada (G-18-0021807).

